# Community Blood Pressure groups – examining the acceptability and other effects of a pilot intervention in Zimbabwe

**DOI:** 10.64898/2026.04.20.26351307

**Authors:** Trevor Chivandire, Ardele Ndanga, Fadzaishe M. Mhino, Cuthbert Sekanevana, Chipo E. Mpandaguta, Takudzwa Mwanza, Alvern Mutengerere, Susana Scott, Pugie Chimberengwa, Justin Dixon, Chiratidzo Ndhlovu, Janet Seeley, Rudo M.S Chingono, Kalpana Sabapathy

## Abstract

**Introduction:** Over one billion people worldwide have hypertension. In Zimbabwe, prevalence is an estimated 38%, surpassing the global average of 34%, and >50% of hypertensives are undiagnosed. The Community BP (Com-BP) groups study examined whether community groups of people living with hypertension, provided with BP machines and led by trained Facilitators could improve awareness, screening and support for those diagnosed with hypertension, to help blood pressure (BP) control. We present findings from the quantitative evaluation of the Com-BP pilot intervention.

**Methods:** The acceptability of the Com-BP group pilot intervention, its potential effectiveness in improving knowledge, attitudes and practices (KAP) and in reducing BP among hypertensive adults in Zimbabwe, was evaluated. Two cross-sectional surveys using standardised questionnaires, and BP and Body Mass Index (BMI) assessments, were done at the start and end of the pilot intervention. Statistical evidence of difference between baseline and follow-up was examined using Wilcoxon signed-rank test for continuous data and McNemar’s test for categorical data.

**Results:** Fourteen groups (seven urban and seven rural) were formed and 151 participants joined over a median of 5months, including 140 enrolled before baseline assessment and 11 who joined during implementation. Retention in the groups was 97.9% (137/140 recruited at baseline), with approximately equal numbers from the urban and rural sites. Median age at baseline was 54 years (IQR 45-66y; min-max 30-92y) and the majority (79%, n=108) were female. Most participants (82.5%, n=113) rated their experience of the group sessions as excellent. The proportions of participants with changes in KAP from baseline to endline were as follows: 45.3% (n=62) to 81.0% (n=111) (p=0.004) able to identify at least two pre-disposing factors for hypertension; 65.0% (n=89) to 77.4% (n=106) (p=0.02) reporting ≥1day of vigorous physical activity/week; 28.5% (n=39) to 13.9% (n=19) (p=0.001) reporting salt added to meals at the table. There was no statistical evidence of any difference in medication adherence, p=0.06. The proportion of participants with uncontrolled hypertension was 57.7 % (n=79) at baseline and reduced to 31.4% (n=43) at follow-up (p<0.001).

**Discussion:** Community groups for improving awareness, detection and support were acceptable and associated with improvements in self-reported KAP and prevalence of uncontrolled BP. Further research on the sustainability and impact of the intervention is required.

## Introduction

Hypertension is the leading modifiable risk factor for cardiovascular morbidity and mortality. It is estimated that approximately 1.4 billion adults worldwide aged between 30 and 79 years are living with hypertension, of whom fewer than 20 per cent. have their condition optimally managed.(1) Prevalence trend analyses involving at least 1200 population-representative studies between 1975 and 2019 showed that while the burden of hypertension is falling in high income countries, it has risen, or at best remained stagnant, in low- and middle-income countries (LMICs) such as those in sub-Saharan Africa.(2,3) In Zimbabwe, hypertension prevalence is an estimated 38%, surpassing the global average of 34%.(1) This high burden is further compounded by health systems that often struggle to deliver sustained hypertension care due to underfunding, resource limitations, and healthcare worker shortages.(4)

In Zimbabwe, an estimated 59% of cardiovascular disease deaths are attributed to hypertension,(1,5) which leads to complications such as stroke, kidney disease, and heart failure.(6,7) Yet, it is estimated that half of those living with hypertension in LMIC settings are unaware of their blood pressure (BP) status.(1,7) Accessible and effective strategies to improve awareness, screening and support for those diagnosed with hypertension could enhance the cascade of care from BP awareness to treatment and optimal control.(1,8)

Community-based interventions, such as those involving community health workers (CHWs), peer support and self-monitoring of blood pressure, are described as key in improving hypertension awareness, treatment adherence and BP control in LMICs by the WHO, including in the HEARTS technical package.(1,9)

The Com-BP groups study was planned to provide pilot data on whether community groups could contribute to improving awareness about BP, knowledge about hypertension and impact multiple steps in the cascade of care for hypertension, while simultaneously empowering individuals to have greater agency for their own health.(10) Evidence on community-based hypertension group models in sub-Saharan Africa remains limited, with existing models generally being health-worker or health-facility led or focused on health promotion without addressing treatment needs.(11,12) Other community-based approaches have focused on task-shifting hypertension care to CHWs, including supporting diagnosis and treatment initiation at the community level.(13) In contrast, the Com-BP groups pilot intervention was strongly community-led. Similar group-based approaches have been successfully implemented in HIV care, where community antiretroviral therapy (ART) refill groups have improved treatment adherence, retention in care, and reduced the burden on health systems.(14,15) Lessons from these community-based HIV care models in high HIV prevalence African settings informed the design of the Com-BP groups intervention.

Key components of the intervention were the use of trained peer community members including a lay CHW to facilitate groups and peer support between members for improved adherence and healthy living, with provision of BP machines for group members and their contacts to measure BP in the community.(10)

Here we present findings from quantitative evaluation of the Com-BP groups pilot intervention to understand the acceptability of the Com-BP groups the scope for improving knowledge, attitudes and practices (KAP), and to explore any BP changes before and after the pilot among hypertensive adults in Zimbabwe.

## Methods

### Ethics Statement

Written informed consent was provided by all participants and anyone unable to write was assisted by a nominated witness. The study was approved by the ethics committees of the Biomedical Research and Training Institute ethics board, Medical Research Council of Zimbabwe (MRCZ A/3069), Research Council of Zimbabwe and London School of Hygiene and Tropical Medicine (Ref: 29785). In addition, relevant authorities in Zimbabwe, including the Ministry of Health and Child Care, the Chitungwiza City Health Department, the Chitungwiza District Administrator and Howard Mission Hospital in Chiweshe provided approval.

### Hypotheses

We hypothesised that Com-BP groups could contribute towards improved knowledge about hypertension and lead to improved control of BP.

### Objectives

The analysis was designed to address three key objectives:

i. To determine the acceptability of the Com-BP groups intervention.
ii. To identify if KAP among group members improved after the pilot intervention (follow-up) compared to at the start of Com-BP groups (baseline).
iii. To identify if mean BP measures were lower at follow-up compared to mean BP measures at baseline.

### Study design

The current analysis presents findings of observational evaluation involving two cross-sectional before and after surveys at baseline and at the end of the Com-BP group pilot intervention respectively. The surveys assessed the acceptability of the intervention at follow-up, and compared KAP and BP measures before and after the Com-BP groups intervention (baseline vs follow-up).

### Study setting and population

The Com-BP groups study was carried out between October 2023 and April 2025 involving three phases of research to co-produce (Phase 1), implement (Phase 2) and evaluate (Phase 3) the Com-BP groups pilot intervention.(10) Participants were recruited and completed baseline assessments between 7 and 30 May 2024. The follow-up survey was between 7 October and 1 November 2024. The study was conducted in an urban (Chitungwiza) and a rural (Chiweshe) community in Zimbabwe. Chitungwiza is a satellite town situated roughly 25 kilometres from the capital city, Harare, and Chiweshe is a rural, agricultural region. In each community, one health facility was selected to serve as the primary referral centre for clinical services.

The current study was part of the second phase of the study, during which Com-BP group members were surveyed at baseline and at the end (follow-up) of the pilot intervention. The surveys involved a standardised questionnaire examining participant characteristics, lifestyle and behaviours, hypertension related medical history, and KAP regarding hypertension and associated adverse outcomes. BP measures, weight and height were done in accordance with World Health Organisation (WHO) guidelines at baseline and follow-up.(16,17) At each time-point, three BP measurements were taken at approximately 1-minute intervals using the appropriate cuff size and a calibrated digital automatic BP monitor (Omron X2 Basic, Omron Healthcare Co., Ltd., Kyoto, Japan). Weight was measured using a calibrated digital scale (Seca; seca GmbH & Co. KG, Hamburg, Germany) and height was measured using a portable stadiometer (Seca 213; seca GmbH & Co. KG, Hamburg, Germany).

The Com-BP groups pilot intervention development is described elsewhere and is summarised in Figure 1.(10) Groups were formed by CHWs working in the area who publicised the intervention and made known that adults aged 18 years and above, who were known to be diagnosed as hypertensive or who were screened with raised BP but not confirmed (with repeat measures by health care worker), were eligible to join. The decision to join was voluntary. Groups were co-facilitated by a local CHW (primary facilitator) and a second co-facilitator who was a member of the group living with hypertension, nominated by the group to co-facilitate. BP machines were provided by the study, for groups to use and share among members and close contacts from their households and neighbours.(10)

**Figure 1:**
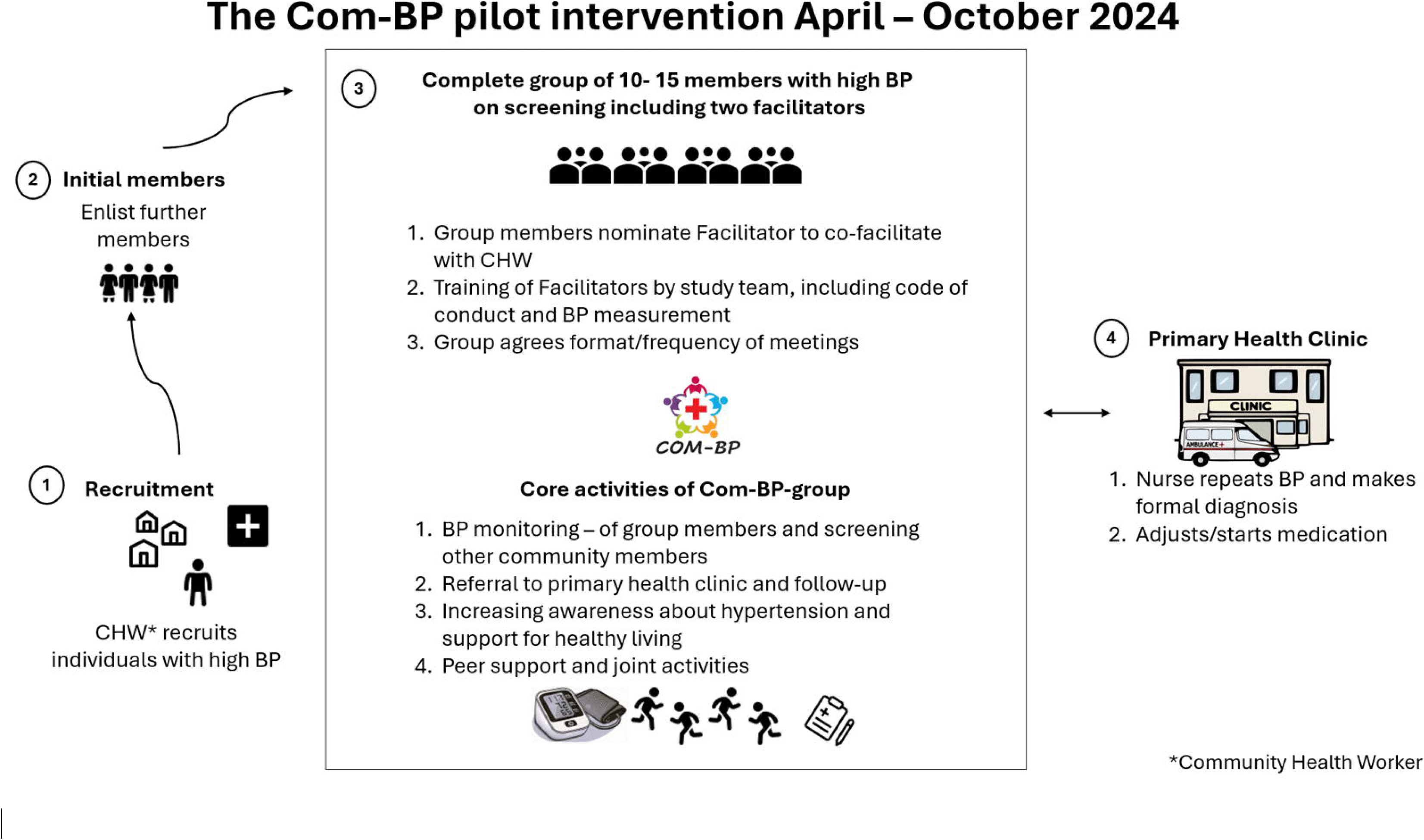
Design of the Community BP groups pilot intervention. The Com-BP intervention involved the formation of community groups of people living with hypertension, facilitated by a community health worker and a nominated group member. Groups were provided with blood pressure monitors and educational materials to support community blood pressure measurement, hypertension education, healthy living and peer support. BP, blood pressure; CHW, community health worker.

Facilitators (CHWs and co-facilitators) received training on hypertension based on a Facilitator’s manual developed in the study. Themes covered BP measurement and interpretation according to WHO guidance, fundamental information and education materials on hypertension, and requirements for an appropriate code of conduct to facilitate the groups. (10,16) They were further supported by standardised materials co-developed as part of the Com-BP groups pilot intervention, including a ‘Hypertension Information’ booklet and film, a topic guide for potential topics to be discussed during meetings, and suggestions for activities related to healthy living e.g. dancing/walking or other exercise (offered as options for the group to choose from).(10) CHWs were responsible for overseeing access to the BP monitors, and community members could check their BP at no cost, with assistance from group facilitators, regardless of whether they were enrolled in the group. CHWs were responsible for referring individuals requiring clinical care.

## Data analysis

### Participant characteristics

We analysed data for participants who completed baseline and follow-up surveys. Age was stratified into four categories (30-45 years, 46-55 years, 56-65 years and ≥66 years) to ensure approximately equal number of participants in each category. Household income was self-reported and analysed using standard categories, and the lowest category further divided to less than US$50 per month and US$50-100 instead of less than US$100 per month due to the high proportion of participants reporting the former. Wealth Index was defined as wealth quartiles using the Kaiser Wealth Index score, constructed through the principal component analysis (PCA) of asset ownership to generate a composite indicator of socioeconomic status (poorest to wealthiest).(18) Assets assessed included ownership of television, radio, microwave, computer/laptop bicycle, refrigerator, stove and mobile phone. Participant’s occupational status was assessed based on the question, “how do you spend most your time during a day?” with response options i). Housework ii). Work from home compound iii). Work or business outside home iv). Education outside home v). Other activity outside home. Historical diagnosis of hypertension was based on participants self-report at the baseline visit.

### Acceptability of the intervention

The acceptability of the intervention was assessed based on the proportion of participants retained and continuing to attend group sessions and participants responses to i) how easy they found it to get to the meeting point; ii) ease of taking time to attend group meetings; iii) participant rating of their experience with attending the groups.

### Changes in Knowledge, Attitudes and Practices (KAP) including medication adherence

Knowledge related change in KAP at follow-up compared to baseline was assessed based on change in knowledge about predisposing factors for hypertension defined as i) ability to identify at least two risk factors of hypertension and ii) change in median score of correctly identifying risk factors for hypertension. Change in the proportion of participants able to identify uncontrolled hypertension as a risk factor for stroke or heart attack was also explored.

In addition, we assessed self-reported median number of days spent doing vigorous physical activity and change in proportion of participants reporting at least one day of such activity. Vigorous physical activity was defined according to the WHO guidance, as engaging in activities such as heavy lifting, digging, aerobics, or fast bicycling at least one day per week. (17) We also examined self-reported use of salt and salty sauces during food preparation and at the table (categorised as rarely, sometimes, often and always).

We assessed willingness and ability to take antihypertensive medication using a 10-item scale for self-reported medication adherence (MAR scale)(19), as no validated tool to measure adherence with hypertensive medication was available in this setting. The scale ranges from 0-10, with 0 indicating high adherence and 10 representing low adherence. High adherence was defined as a total score of between 0-2, moderate adherence between 2-4 and lower adherence as ≥5. We further reported median self-reported adherence levels at follow-up compared to baseline.

### Changes in clinical characteristics

Of the three BP measures done for each participant at each time point (baseline and follow-up), the average of the last two systolic (SBP) and last two diastolic BPs (DBP) measures were calculated to estimate an individual’s BP based on the WHO STEPS protocol.(17) BP was categorised according to International Society of Hypertension guidelines as follows: uncontrolled hypertension as SBP greater than or equal to 140 mmHg, DBP greater than or equal to 90 mmHg, or both; high-normal BP as SBP 130–139 mmHg, DBP 85–89 mmHg, or both; and normal BP as a SBP less than 130mmHg and DBP less than 85 mmHg.(20)

Body mass index (BMI) was calculated as weight in kilograms divided by the square of height in metres (kg/m^2^) and categorised according to WHO guidance as follows; i) < 18.5 kg/m^2^, underweight; ii) 18.5 - 24.9 kg/m^2^, normal weight; iii) 25.0 - 29.9 kg/m^2^, overweight; and iv) ≥ 30 kg/m^2^, obesity.(21,22)

BP change was defined as clinically significant if there was a change in systolic BP of ≥10mmHg in a given participant’s BP between baseline and follow-up measurements, based on evidence demonstrating that a 10mmHg reduction is associated with approximately 20% lower risk of major cardiovascular events.(23) We also assessed changes in mean population BP at follow-up compared to baseline.

### Statistical considerations

Categorical variables were reported as percentages (%) and frequencies (n), and continuous variables as median and interquartile range (IQR). At baseline, we assessed differences between categorical variables using the Pearson chi-square test or Fisher’s exact test where expected cell counts were under five. Comparison of participant characteristics and acceptability of the intervention were made by sex and community. Evidence of difference in KAP and BP between baseline and follow-up were examined using the Wilcoxon signed-rank tests for continuous and ordinal data and the McNemar’s tests for binary categorical data. For all tests, statistical significance was considered if p-values ≤0.05. All analyses were performed using STATA software version 13.1 (Stata Corporation College Station, TX). Data visualisations were performed R Studio version 4.3.0.

The sample size of 140 participants which the was the target for two sites, was based on feasibility of conducting approximately 7 groups consisting of 10 individuals per group at the urban and rural sites, respectively. Group size was agreed with participants during formative work (co-development, Phase 1) of the pilot Com-BP groups intervention.(10) A sample of 140 participants estimates the true population proportion if acceptability is 90% with 95% CI 84%-94%. While the primary objective of this study was acceptability, secondary analyses evaluated changes in clinical and behavioural outcomes, including SBP and KAP differences by community. The study was not powered to detect specific effect sizes for these secondary clinical endpoints and we may be under-powered to detect more subtle differences.

## Results

### Participant characteristics at baseline

The Com-BP groups pilot intervention was conducted for a mean duration of five months and is summarised below in Figure 2.

**Figure 2:**
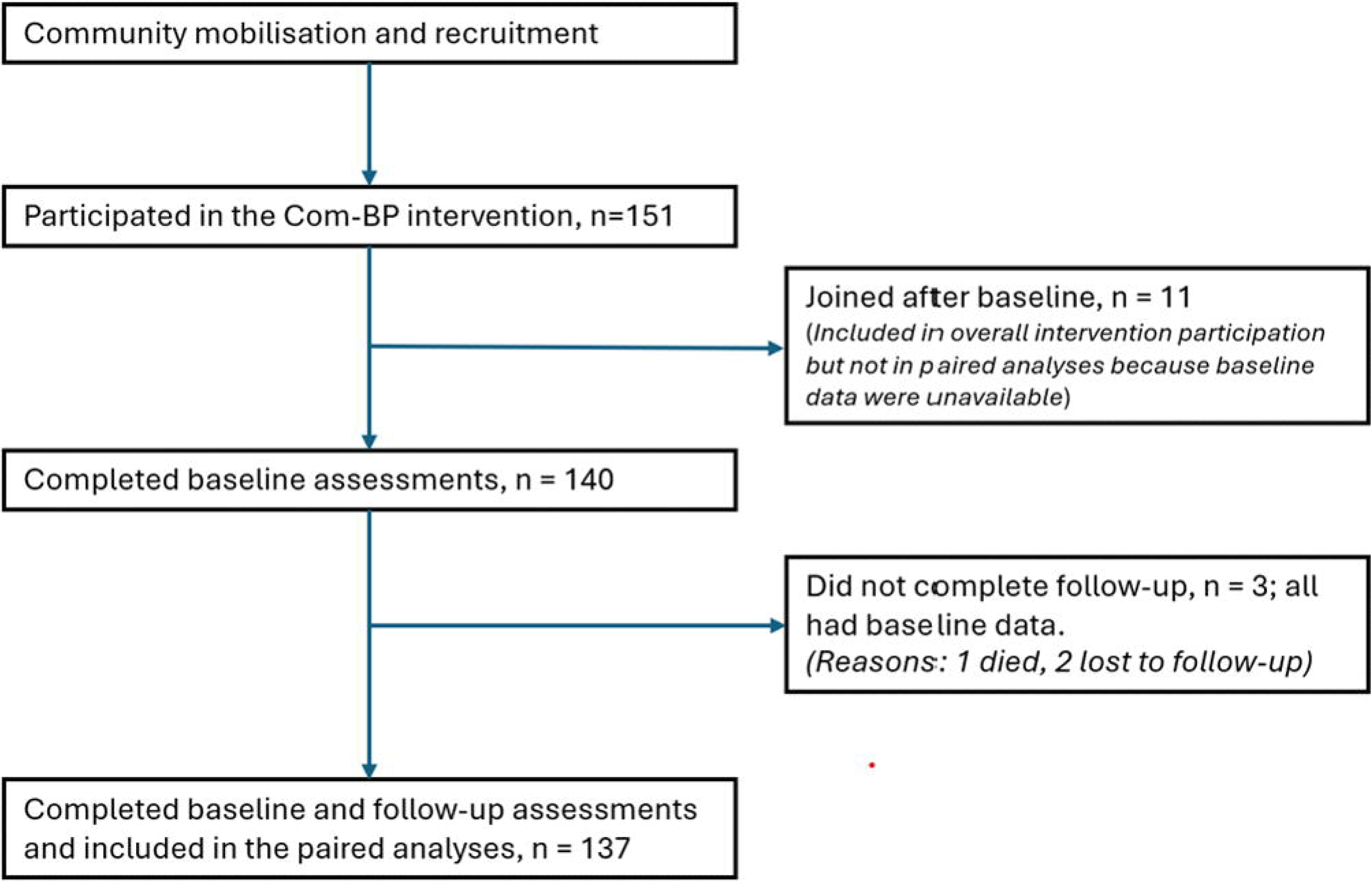
Participant flow in the Com-BP groups intervention. A total of 151 participants joined the Com-BP groups during the intervention period. Of the 140 participants recruited at baseline, 137 completed the follow-up assessment and were included in the paired analysis. Eleven participants joined after baseline assessments and were not included in the analysis because baseline measurements were unavailable.

There were 140 participants recruited at baseline and a further eleven joined the groups during the intervention period. Three baseline participants did not complete the follow-up survey. As such, almost 98% of baseline participants completed baseline and follow-up surveys (137/140, 97.9%) and approximately equal numbers were from the urban (51.1%, n=70) and rural sites (48.9%, n=67) (Table 1). Median age at baseline was 54 years (IQR 45 – 66) with an age range of 30 – 92 years, and the majority (78.8%, n=108) were female. For men, the median age was age was 59 years (IQR 52 – 70), and it was 53 years (IQR 44 – 65) for women, p=0.38. Overall, 32.1% (n=44) participants were widowed, 38.9% (42/108) of women vs 6.9% (2/29) of men, p=0.005. Of the participants, 61.3%, n=84 had received up to secondary/post-secondary education. More participants from urban compared to rural community had completed secondary/post-secondary education (81.4% (57/70) vs 40.3% (27/67), p <0.001.

**Table 1:**
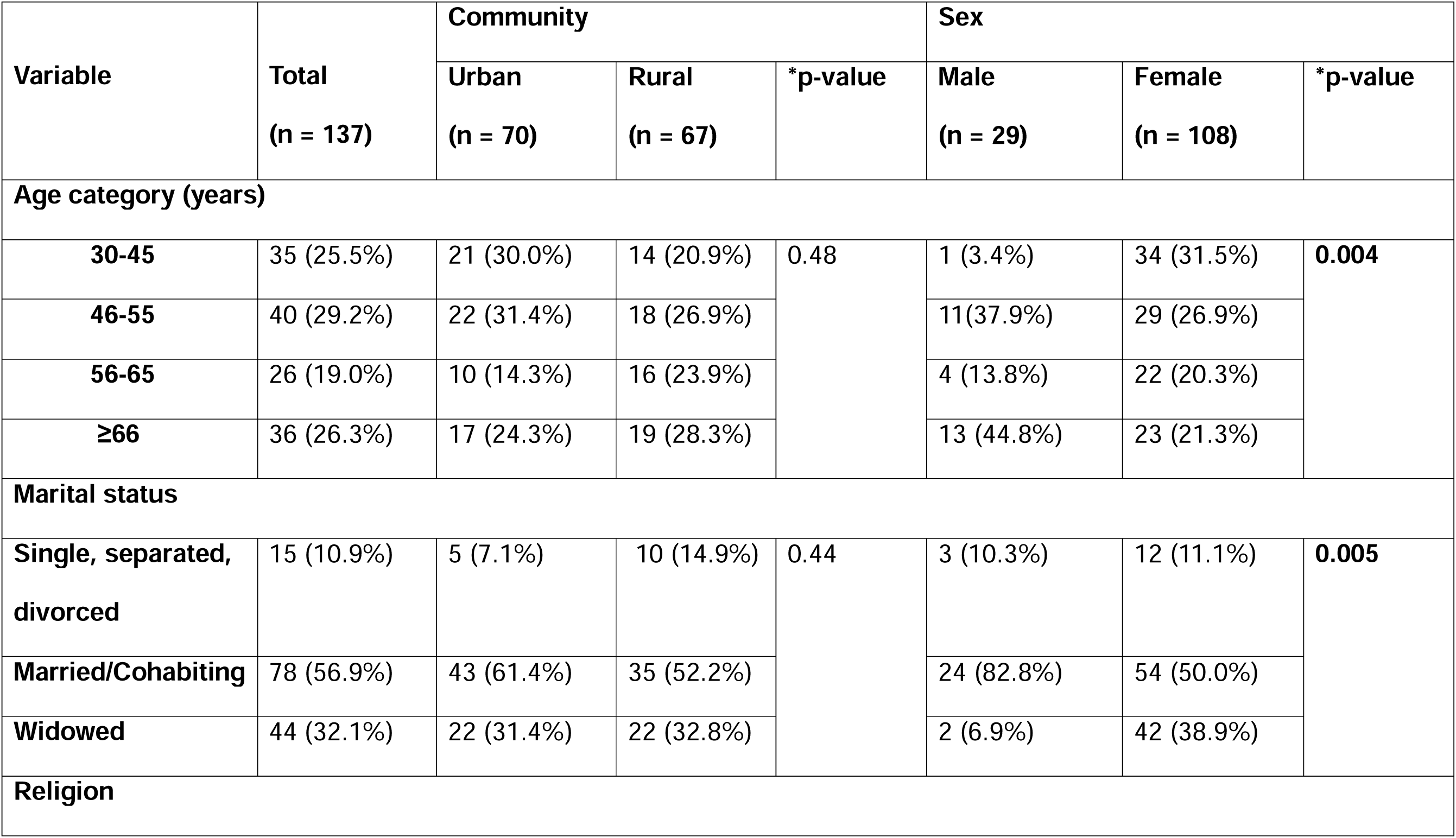
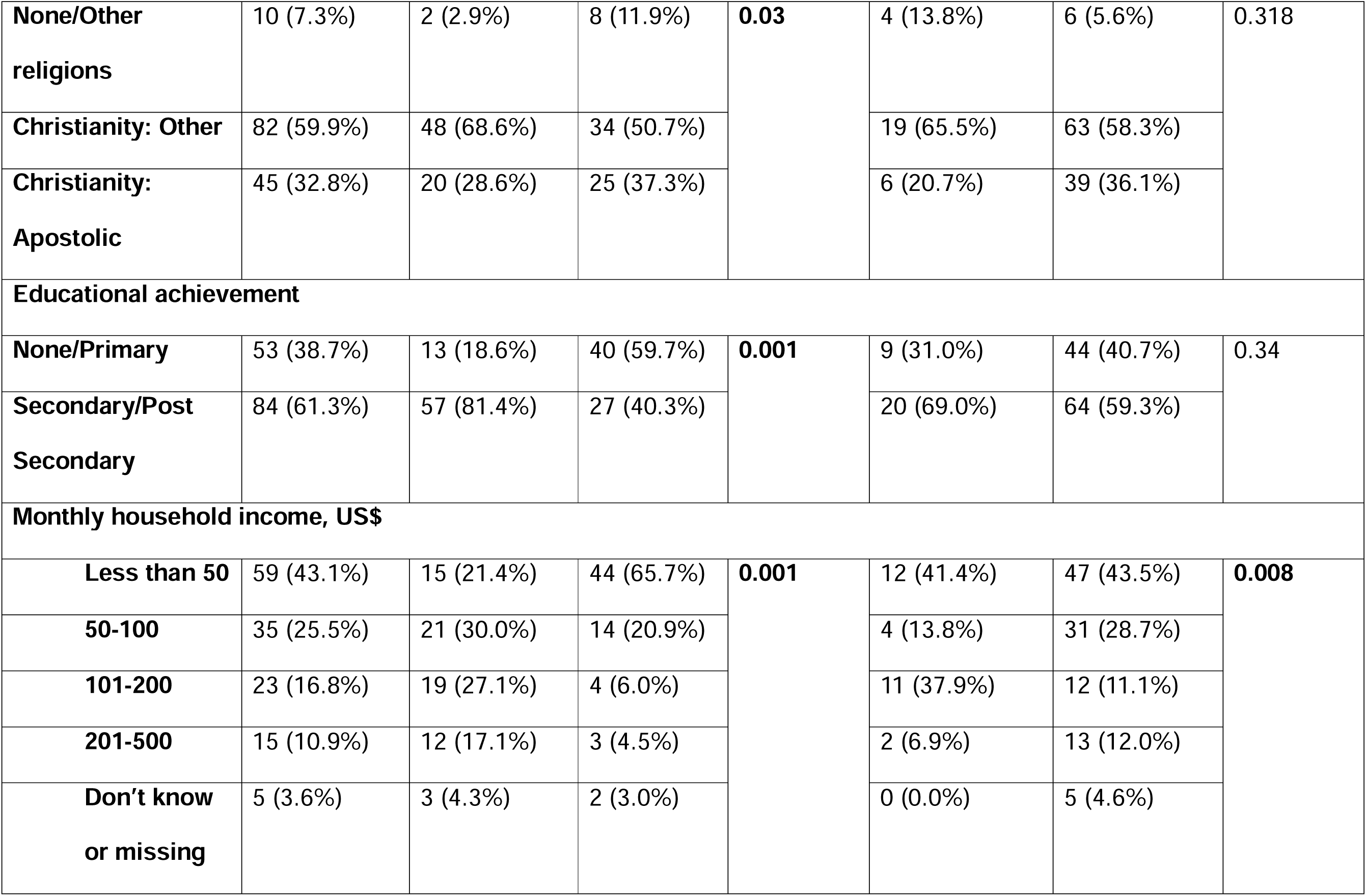
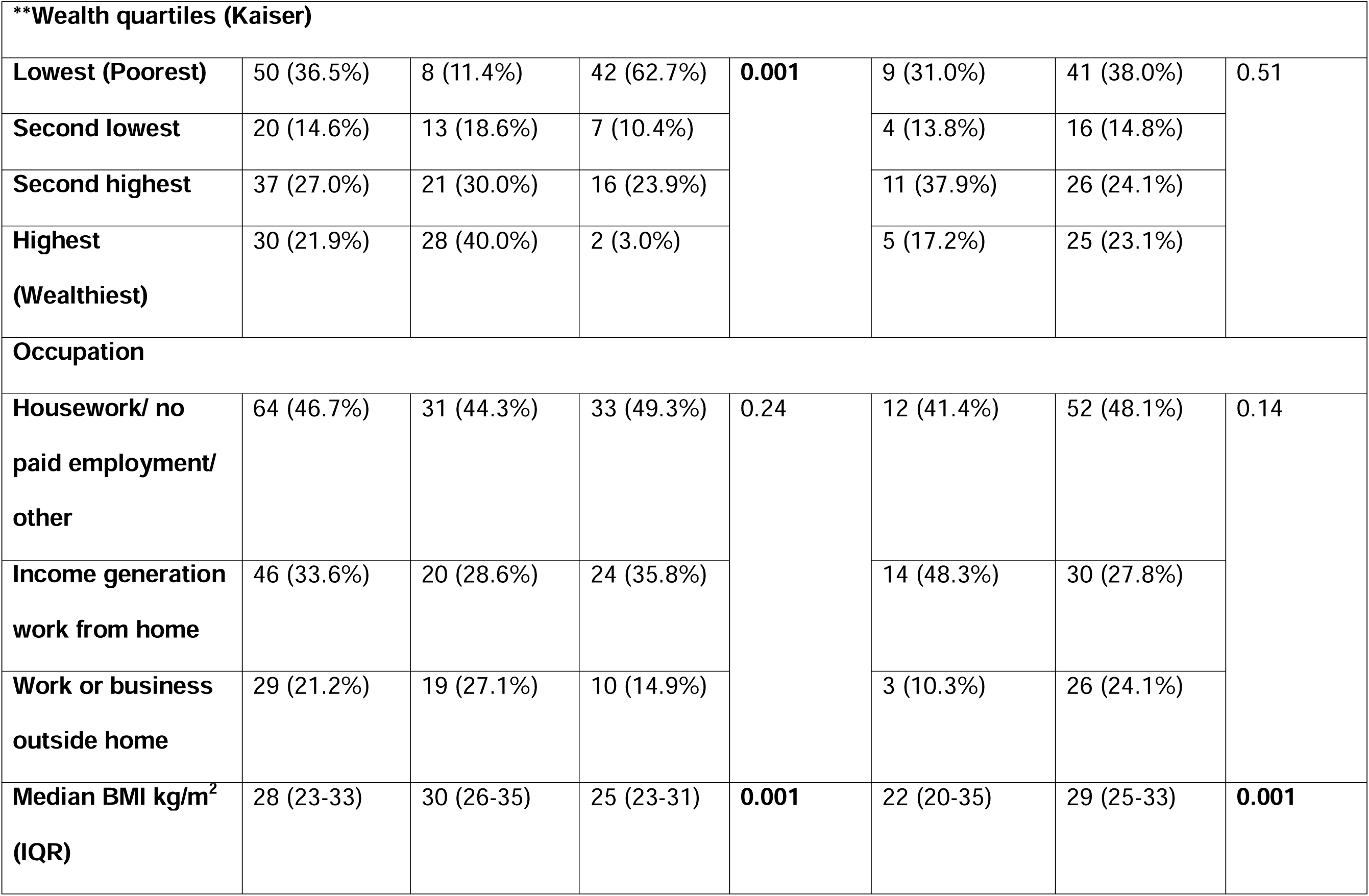
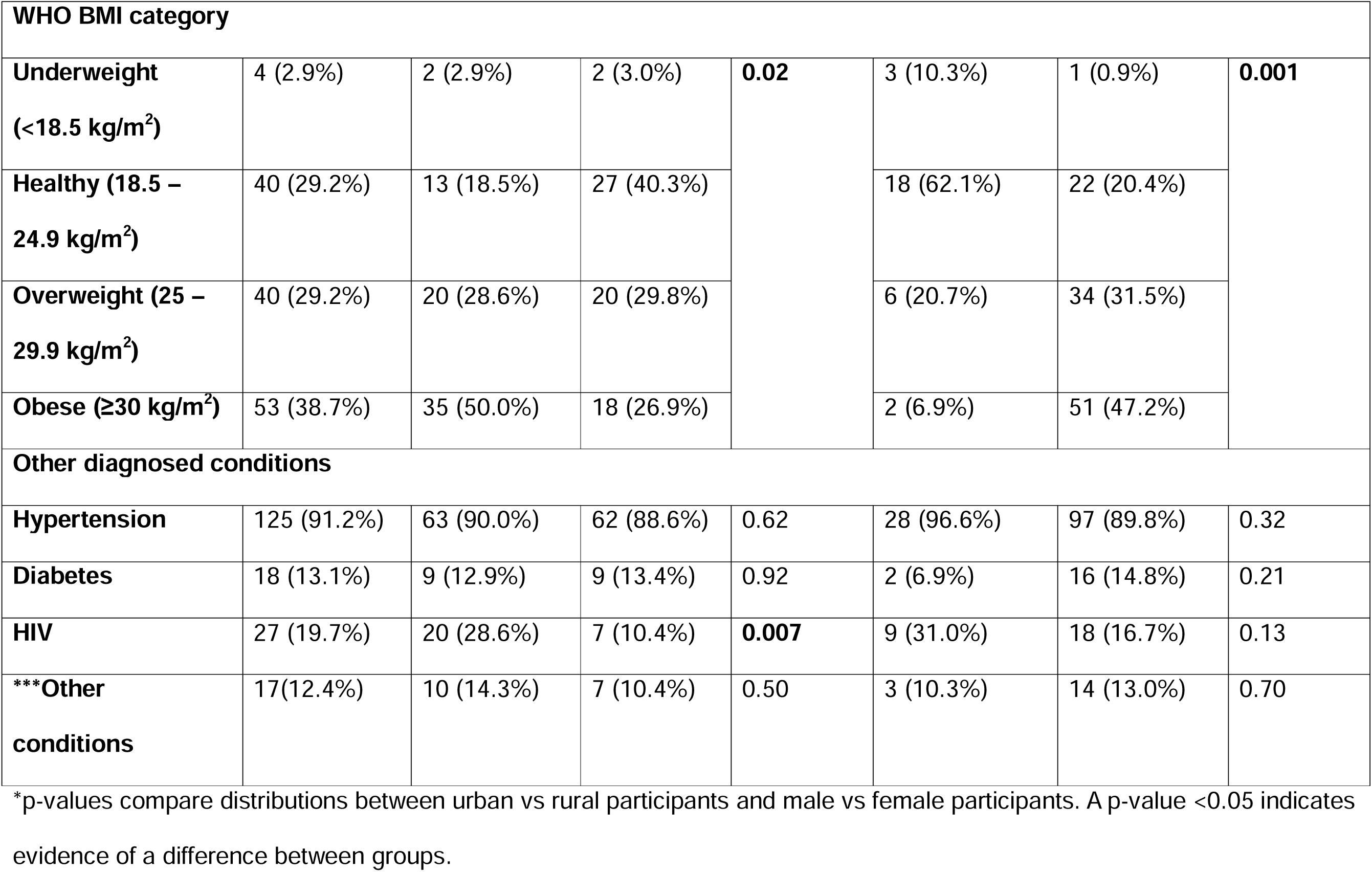
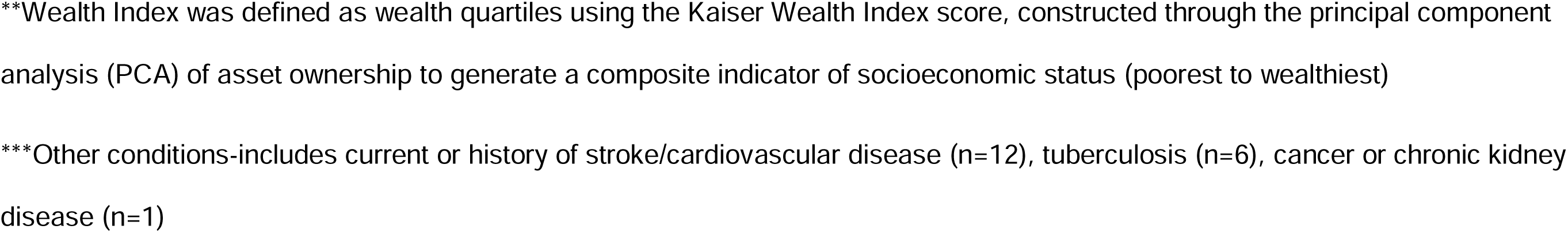
Baseline sociodemographic characteristics of participants stratified by community and sex

A high proportion (43.1%, 59/137) of participants reported a monthly household income of less than US$50 and in the rural community this was the case for the majority (65.7% (44/67) vs 21.4% (15/70) in the urban community, p<0.001). Forty percent (28/70) of urban community members reported asset ownership which placed them in the wealthiest wealth quartile for the study population, compared with 3.0% (2/67) among rural community members, p<0.001. The majority of the latter were in the poorest quartile (62.7%, 42/67). The Christian apostolic church was the largest single faith group (32.8% (45/137) irrespective of community, with other participants reporting several other Christian denominations.

Median body mass index (BMI) was 28 kg/m^2^ (IQR 23 – 33). In the urban community, median BMI was 30 kg/m^2^ (IQR, 26-35) compared to 25 kg/m^2^ (IQR, 23 – 30) in the rural community, p<0.001. Median baseline BMI in males was 22 kg/m^2^ (IQR 20 – 25) and 29 kg/m^2^ (IQR 25-33) in females, p<0.001.

Ninety one percent (125/137) of the participants had a diagnosis of hypertension before enrolment into the cohort (89% (111/125) taking antihypertensive medication). Diabetes and living with HIV were the other two most reported co-morbidities (13.1% and 19.7%, respectively), with the latter being more frequently self-reported by urban than rural participants (28.6% versus 10.4%, p=0.007).

### Acceptability of the intervention

Acceptability based on retention and continued attendance was high with 97.9% (137/140) of participants recruited at baseline (n=140) present during the follow-up survey. The majority of participants rated their experience of the group sessions to be excellent (82.5%, n=113), with 94.9% (130/137) reporting it easy to take time off their schedules and attend group sessions. Most participants were satisfied (84.7%, 116/137) with having access to the BP measurement at the community level (Table 2). The intervention was equally acceptable among members attending urban and rural groups, and men and women.

**Table 2:** Acceptability of the Com-BP groups intervention, stratified by community and sex

| Variable | Overall<br>(n=137) | Urban<br>(n=70) | Rural<br>(n=67) | *p-value | Male<br>(n=29) | Female<br>(n=108) | *p-value |
| --- | --- | --- | --- | --- | --- | --- | --- |
| i) Ease of getting to the meeting point (easy/very easy) | 130 (94.9%) | 67 (95.7%) | 63 (94.0%) | 0.13 | 27 (93.1%) | 103 (95.4%) | 0.42 |
| ii) Ease of taking time to attend session ((easy/very easy)) | 130 (94.9%) | 64 (91.4%) | 66 (98.5%) | 0.14 | 27 (93.1%) | 103 (95.4%) | 0.36 |
| iii) Overall rating of group as excellent | 113 (82.5%) | 53 (75.7%) | 60 (89.6%) | 0.08 | 21 (72.4%) | 92 (85.2%) | 0.28 |

\*p-values compare distributions between urban vs rural participants and male vs female participants. A p-value <0.05 indicates evidence of a difference between groups.

### Knowledge, attitude and practices at baseline and follow-up

The change in participants’ knowledge of the risk factors for hypertension: alcohol consumption, lack of physical activity, cholesterol, salt consumption, stress, sugar consumption and obesity, is shown in Table 3. Participants identified a median of one (IQR 1-3) risk factor for hypertension at baseline, and this rose to a median of three (IQR 2 – 4) at follow-up, p <0.001. The proportion of participants who could identify at least two pre-disposing factors for hypertension increased from 45.3% (62/137) at baseline to 81.0% (111/137) at follow-up (p=0.004). There was an increase in the proportion of participants correctly identifying all risk factors except stress, which was recognised as a predictor by 95% of participants at baseline and endline (Table 3).

**Table 3:** Change in knowledge related to hypertension from baseline to follow-up

| Variable | Baseline, n (%) | Follow-up, n (%) | *p-value |
| --- | --- | --- | --- |
| Median (IQR), number of risk factors known | 1 (1-3) | 3 (2-4) | **<0.001** |
| **Number of risk factors** |  |  |  |
| **0 – 1** | 76 (55.4%) | 26 (19.0%) | **<0.001** |
| **2** | 20 (14.6%) | 31 (22.6%) |  |
| **3** | 28 (20.4%) | 36 (26.3%) |  |
| 4 | 11 (8.0%) | 15 (10.9%) |  |
| 5 or more | 5 (3.6%) | 29 (21.2%) |  |
| Factors accurately known as risk factors for hypertension |  |  |  |
| Overweight/obesity | 24 (17.5%) | 47 (34.3%) | 0.002 |
| Lack of physical activity | 16 (11.7%) | 40 (29.2%) | <0.001 |
| Alcohol consumption | 3 (2.2%) | 36 (26.3%) | <0.001 |
| Excess salt consumption | 55 (40.1%) | 101 (73.7%) | <0.001 |
| Stress | 133 (97.1%) | 130 (94.9%) | 1.00 |
| Cholesterol as a risk factor | 31 (22.6%) | 52 (37.9%) | 0.005 |
| Vigorous physical activity | 89 (65.0%) | 106 (77.4%) | 0.02 |
| Salt added at table | 39 (28.5%) | 19 (13.9%) | 0.001 |
| Salt/salty sauce usage at food preparation | 129 (94.2%) | 121 (88.3%) | 0.09 |

\*p-values compare distributions between baseline and follow-up. A p-value <0.05 indicates evidence of statistical difference between groups. For ordinal data, p-values are from a single test evaluating the evidence for a difference overall between groups in the distribution of outcomes across categories of the given variable.

At baseline 57.7% (79/137) identified uncontrolled hypertension as a risk factor for stroke, and the proportion at follow-up was 73.7% (101/137), p=0.01. The proportion of participants who could identify uncontrolled hypertension as a risk factor for heart attack at baseline was 38.0% (52/137) versus 61.3% (84/137) at follow-up, p <0.001.

There was no difference in the proportion of participants who perceived that they were at risk of stroke/heart disease between baseline and endline (from 73.7% (101/137) to 75.9% (104/137), p=0.08). The proportion of participants reporting at least one day of vigorous physical activity per week was 65.0% (89/137) at baseline vs 77.4% (106/137) at follow-up, p=0.02. Overall, 28.5% (39/137) of participants reported adding salt to meals at the table, at baseline vs 13.9% (19/137)) at follow-up, p=0.001. There was no statistical significance of any difference in medication adherence as measured by the medication adherence reporting scale when comparing baseline with follow-up (p=0.06).

### BP at baseline and follow-up

Median SBP at baseline was 135 mmHg (IQR 125 – 153), and median DBP was 89 mmHg (IQR 82 – 97). At follow-up, approximately 5 months later, median SBP was 127 mmHg (IQR 116 – 136) and median DBP was 84 mmHg (IQR 77 – 90). The proportion of participants with uncontrolled hypertension (i.e. change in BP category from <140/90 mmHg to ≥140/90 mmHg) decreased from 57.7% (n=79) at baseline to 31.4% (n=43) at follow-up (p<0.001), and the proportion with BP in normal range (<130/85 mmHg) increased from 25.5% to 45.3% (p<0.001) (Table 4).

**Table 4:** Change in BP from baseline to follow-up

| Variable | Baseline | Follow-up | *p-value |
| --- | --- | --- | --- |
| Median SBP/mmHg (IQR) | 135 (125-153) | 127 (116-136) | **<0.001** |
| Median DBP/mmHg (IQR) | 89 (82-97) | 84 (77-90) | **<0.001** |
| **\*\*BP category, n (% , 95% CI)** |  |  |  |
| Normal BP | 35 (25.5, 18.9-33.6) | 62 (45.3, 37.0-53.8) | **<0.001** |
| High-normal BP | 23 (16.8, 11.4-24.1) | 32 (23.4, 17.0-31.3) |  |
| Hypertension | 79 (57.7, 49.1-65.8) | 43 (31.4, 24.1-39.7) |  |

\*p-values compare distributions between baseline and follow-up. A p-value <0.05 indicates evidence of a difference between groups.
\*\*Normal BP is SBP<130mmHg and DBP<85mmHg; High-normal BP is SBP of 130-139mmHg and/or DBP of 85-89mmHg; Hypertension is SBP≥140mmHg and/or DBP≥90 mmHg.

A clinically significant decrease in SBP (by ≥10 mmHg) was observed in 54.0% (74/137) of participants at follow-up. The overall mean change in SBP was -9.2 mmHg (95%CI -12.2 to -6.16) (Figure 3). There was no evidence of differences in BP changes between baseline and endline by sex ((p=0.07) and by community (p=0.53).

**Figure 3:**
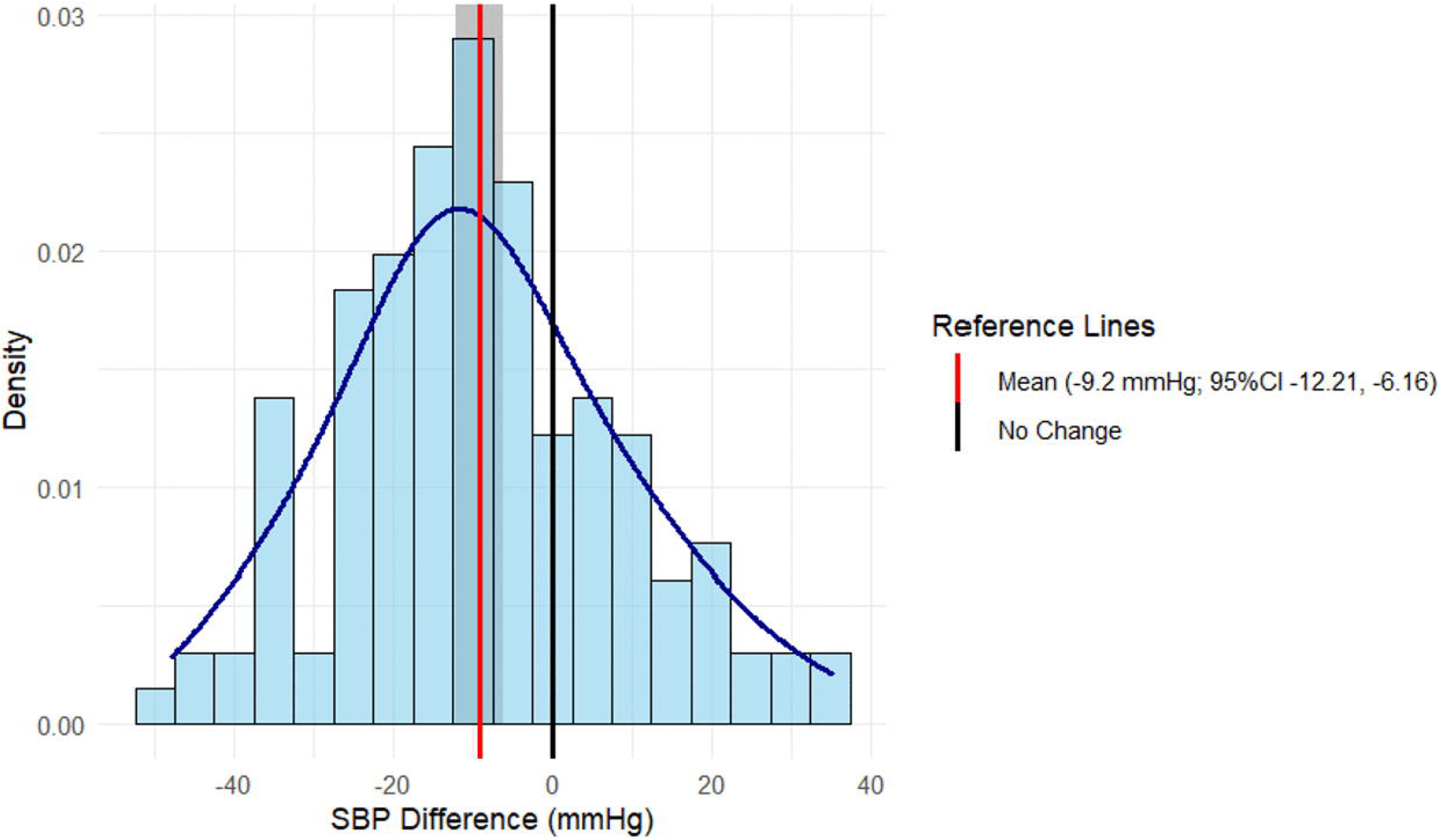
Within-individual mean SBP change between baseline and follow-up. The figure shows the distribution of within-participant change in systolic blood pressure (SBP) between baseline and follow-up. Negative values represent a reduction in SBP at follow-up. The red vertical line indicates the mean change in SBP (−9.2 mmHg; 95% CI −12.21 to −6.16), while the black vertical line indicates no change. CI, confidence interval.

### BMI at baseline and endline

Median BMI at baseline was 29.3 kg/m^2^ (IQR 25.5 - 33.4) and 29.3 kg/m^2^ (IQR 25.1 - 32.9) at follow-up, with no statistical evidence of difference in median BMI despite the majority of participants demonstrating a reduction in their BMI (55.5%, 76/137).

## Discussion

The Com-BP groups pilot intervention was found to be highly acceptable with high retention over approximately 5 months of observation and positive responses about attending the groups. Notably, the proportion of participants with uncontrolled hypertension, despite the majority already taking treatment at baseline, was almost halved follow-up (31% vs 58% at baseline). Knowledge of hypertension and self-reported behaviours were also higher.

Given the voluntary nature of joining Com-BP groups, the characteristics of group members may be indicative of sub-groups of the community for whom the group intervention is well suited. Com-BP groups were formed of community members with hypertension or those not formally diagnosed but with the suspicion that they had an elevated BP based on previous measurement. The participants in our study were predominantly female (78.8%), with a median age of 54 years. Fewer younger adults took up the intervention, especially young men, with only one man aged less than 45 years participating in the groups. Hypertension is perceived to be a condition which affects older people only and fewer younger people would have been aware that they have hypertension or are at risk, and this in turn is reflected in those who joined the groups.(10) These findings limit the generalisability of the results to younger men and, more broadly, to sub-sets of the population who are like the individuals who chose not to participate in the intervention.

Poverty was common and worse among participants from the rural site, likely reflecting the underlying socio-economic status of the communities involved in the study, with rural communities being even poorer in Zimbabwe than in urban areas.(24) It may also reflect the fact that wealthier community members were less inclined to join the groups. Among those taking part in the groups, despite some differences in participant characteristics, our data suggest that acceptability is equally high in urban and rural areas, and among men and women.

Knowledge about risk factors leading to hypertension, and about the role of hypertension in contributing to stroke and heart attack, was higher at follow-up than at baseline. The reduction in salt added at the table, contrasted with the trend in salt used during food preparation, although the statistical evidence for the latter was weak. This may have been due to inconsistencies in reporting, lack of improvement in awareness by the individual cooking the food if the participant was not the one cooking the food, or suggest that individual, conscious dietary choices are more readily modifiable than cooking habits for food that is consumed collectively in families.

Our findings suggest that our group model, involving joint facilitation by a lay CHW and a group member who are appropriately trained and take on the role of “community champion”, can be a way to improve awareness about hypertension and associated conditions. This is consistent with findings from a systematic review of community-based interventions in LMICs that showed improvements in cardiovascular disease risk factor knowledge and health-related behaviours, particularly diet and physical activity.(25) Given that low awareness has been identified as an important barrier in hypertension control in Southern Africa, Com-BP groups may be an important addition to existing means of health promotion.(1,26)

A further notable finding of this study is the lower population mean BP and proportion of participants with uncontrolled BP at follow-up than at baseline, with more than half of the study sample having a SBP which was ≥10 mmHg lower than the SBP at baseline. Potential explanations for this include improvements in lifestyle and medication adherence. Participants reported improvements in lifestyle, although this must be interpreted considering social desirability bias. However, the observed BP reductions should be interpreted cautiously. Regression to the mean is an important consideration in studies measuring BP repeatedly, particularly where participants may have entered the intervention following an elevated BP measurement.(27) Seasonal variation may also have influenced the findings, as BP is generally lower during warmer periods than during colder periods, and seasonality was not accounted for because BP change was a secondary outcome.(28) In addition, a Hawthorne effect is possible, whereby participants modified their behaviour because they were aware that they were being observed.(29)

While weight loss was not a pre-specified objective of the study, it could be an objective indicator of lifestyle improvements. We did not find statistical evidence in reduction of median BMI overall, but among female participants who made up the majority of the sample, there was statistical evidence of difference in median BMI from baseline to follow-up, with 58.3% (63/108) having a lower BMI at follow-up. No such significant difference in median BMI was detectable among men, but only 8/29 men (27.6%) were overweight or obese at baseline, compared with 85/108 women (78.7%) (Table 1). Similarly, in participants from the urban site where overweight/obese was more prevalent, there was statistical evidence of decrease in BMI from baseline to follow-up, whereas no change in BMI was observed in the rural site (Supplementary Table S1). Our findings suggest that further research with a larger sample to increase power and to allow for multivariable analysis to adjust for potential confounders is warranted to examine whether a group intervention could help with weight reduction among overweight or obese individuals in whom weight loss would be beneficial. This in turn could help lower BP and reduce risk of other related chronic conditions including diabetes and heart disease.

During in-depth interviews and focus group discussions, participants reported adhering better to medication after acquiring education regarding the importance of taking medication as prescribed,(10) but we did not identify any quantitative difference when measured by the Medication Adherence Report Scale.(19) This tool has not been validated in contexts similar to our study setting in Zimbabwe and the findings should be interpreted in that light.(25) In contrast, the CLUBMEDS study in Nigeria used a Visual Analogue Scale (VAS) and reported improvements in adherence and concomitant blood pressure reduction.(30) This is consistent with improvements described by our participants during process evaluation of the Com-BP groups intervention.(10) Further research is needed on the optimal tool for examining adherence to hypertension and other NCD medication in the southern African setting.

Peer support groups for people living with HIV are an established model to improve anti-retroviral treatment adherence and management of HIV, and are recommended by the WHO.(31,32) Evidence on the use of the community groups in LMICs for NCDs including hypertension is more limited.(33,34) While this is the case, important exceptions exist. In rural Zimbabwe, Chimberengwa *et al.* demonstrated clear clinical benefits through cooperative inquiry groups and village health worker (VHW)-led hypertension clubs, achieving treatment compliance and stable blood pressure control among participants.(35) Otieno *et al* in Kenya and Ghana demonstrated improvements in uncontrolled hypertension, just as we did, with an intervention that was delivered over one year and included support with advanced technologies such as telemedicine and remote monitoring and management by clinicians.(33) This suggests that the Com-BP groups intervention may have potential in resource-limited contexts, as BP improvements were observed despite its shorter duration and less technology-intensive, predominantly lay-person led model. Peer support and peer-to-peer learning within such group models may act as key mechanisms for behaviour change, through shared experiences, reinforcement of health messages, and collective accountability, which may also be key in the improvements observed in these interventions.(33–35)

### Strengths and limitations of study

The underpinning strength of the Com-BP groups study was the intervention which was developed with close involvement of key stakeholders. This led to multiple strengths of the intervention which are described in detail in the paper by Mhino/Chingono *et al*.(10) These included flexibility of choice of timing and location of meetings and access to health facility enabled by close links with CHW yet not constrained by health-facility imposed fixed requirements which are faced by facility-led groups. This led to the other key strength of the study which was minimum loss to follow up. The high level of community-ownership and self-enablement within the intervention, which involved the use of peers and community health workers already known within the participating communities meant that engagement was sustained. A further strength of the Com-BP groups study was the focus on the first step of the cascade of care, namely improving knowledge about BP and awareness of BP status (BP measure, controlled or uncontrolled hypertension etc) by screening in the community. Addressing the first step has the potential to unlock further steps along the cascade including seeking care and adherence with treatment and compliance with medical advice, having gained improved understanding of the importance of doing so.

The limitations include not having the resources to improve staff knowledge and skills with respect to hypertension management, nor to address a key barrier to on-going sustained BP control caused by cost related barriers to access care and medication. As has already been discussed, the self-selecting nature of the participants who chose to be involved in Com-BP groups, would also impact the high levels of acceptability of the intervention. It is likely that other sub-groups in the community may not find a group model helpful, for example those with work commitments or personal reasons to prefer individualised care. As such, the Com-BP groups intervention should be regarded as one potential addition to improve awareness and screening for some sub-groups in the community, while others may require other approaches. The pilot nature of the study and the before and after comparison using two cross-sectional surveys limits causal inference, as observed changes may have been influenced by regression to the mean, seasonal variation and Hawthorne effects.

The sample size was powered for the primary acceptability objective. The study was, however, not powered a priori to detect specific effect sizes or subgroup differences in secondary clinical and behavioural outcomes, as BP and KAP changes between communities. The study may therefore have been underpowered to detect subtle differences in these outcomes. Analyses were conducted at the individual level rather than adjusting for group-level clustering, as our limited number of clusters (14 groups, 137 participants) precluded the reliable use of advanced clustered modelling techniques. Consequently, the shared within-group dynamics may have influenced standard error estimates, meaning our reported significance levels should be viewed cautiously. Additionally, given the exploratory nature of this pilot study, no adjustment was made for multiple comparisons, increasing the possibility that some statistically significant findings may have occurred by chance (Type 1 error).

### Conclusions and Recommendations

Our study provides preliminary evidence that Com-BP groups may support the first step in the hypertension cascade of care. There were also observed improvements in other clinical measures, which require further investigation in a larger study with longer duration of follow-up. To comprehensively address the full cascade of care for people living with hypertension and associated chronic conditions – from improving detection, to monitoring and treatment initiation, to sustained optimal control long-term – further research of a comprehensive package of care is warranted. Priorities include examining the requirements for scale-up of Com-BP groups to achieve their full potential in Zimbabwe and other southern African settings; approaches to strengthen the capacity of healthcare workers to manage hypertension and associated chronic conditions (including the potential contribution from lay CHWs and “community champions”); and innovations to address financial constraints of long-term treatment and care.

## Data Availability

All data produced in the present study are available upon reasonable request to the authors.

## Acknowledgements

We acknowledge the contribution of all key stakeholders involved in design of the intervention and especially thank community members for their participation in the research.

## Data Availability

The data underlying the findings are available on the London School of Hygiene and Tropical Medicine Data Compass repository upon publication (LSHTM Data Compass).

## Supporting Information

**Supplementary Figure 1: Sex stratified within-individual mean SBP change between baseline and follow-up.** The figure shows the distribution of within-participant change in systolic blood pressure (SBP) between baseline and follow-up, stratified by sex. Negative values represent a reduction in SBP at follow-up. The red vertical lines indicate the mean change in SBP for each group, while the black vertical lines indicate no change. CI, confidence interval.

**Supplementary Figure 2: Community within individual mean SBP change between baseline and follow up.** The figure shows the distribution of within-participant change in systolic blood pressure (SBP) between baseline and follow-up for participants from the urban and rural sites. Negative values represent a reduction in SBP at follow-up. The red vertical lines indicate the mean change in SBP for each group, while the black vertical lines indicate no change. CI, confidence interval.

**Supplementary Table 1: Change in BMI from baseline to follow-up, overall and by residential area and sex.** BMI, body mass index. P-values compare paired BMI measurements at baseline and follow-up using the Wilcoxon signed-rank test.

