## Supplementary figures and images for "Community Blood Pressure groups – examining the acceptability and other effects of a pilot intervention in Zimbabwe"

### Supplementary Figure 1

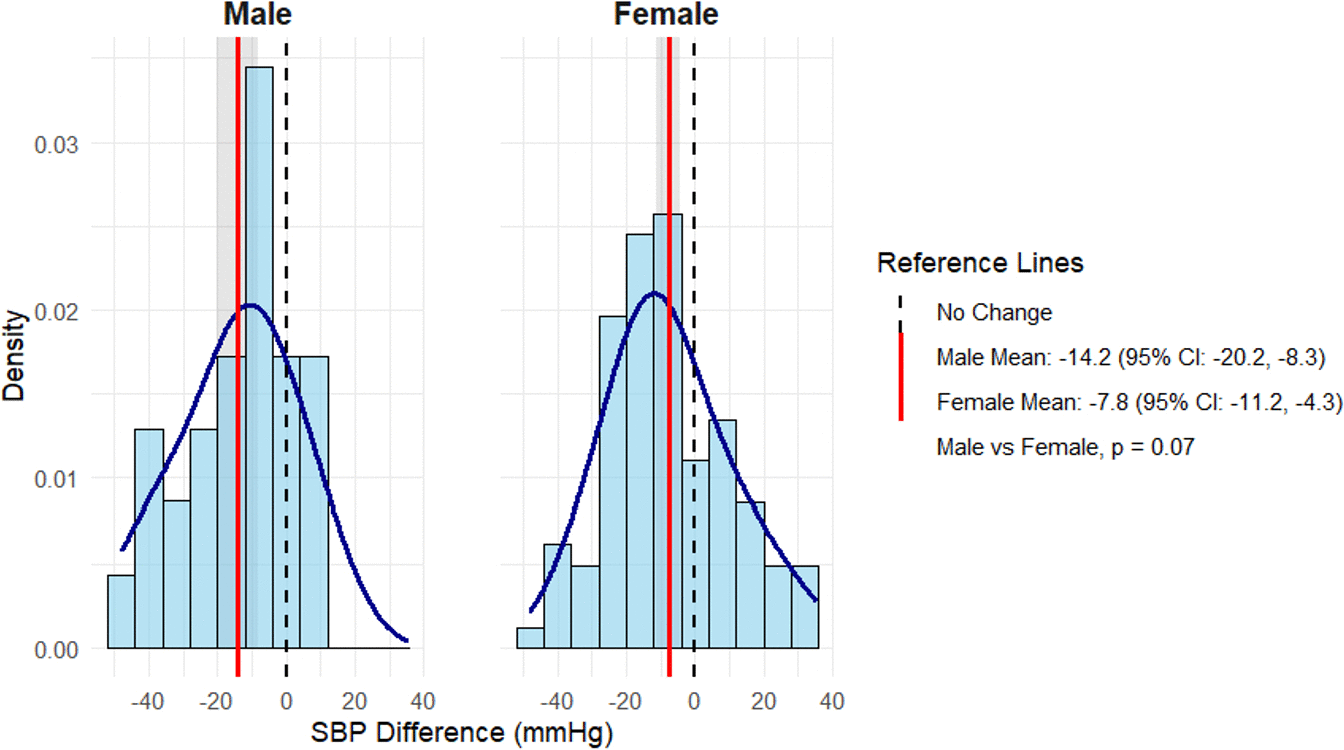

### Supplementary Figure 2

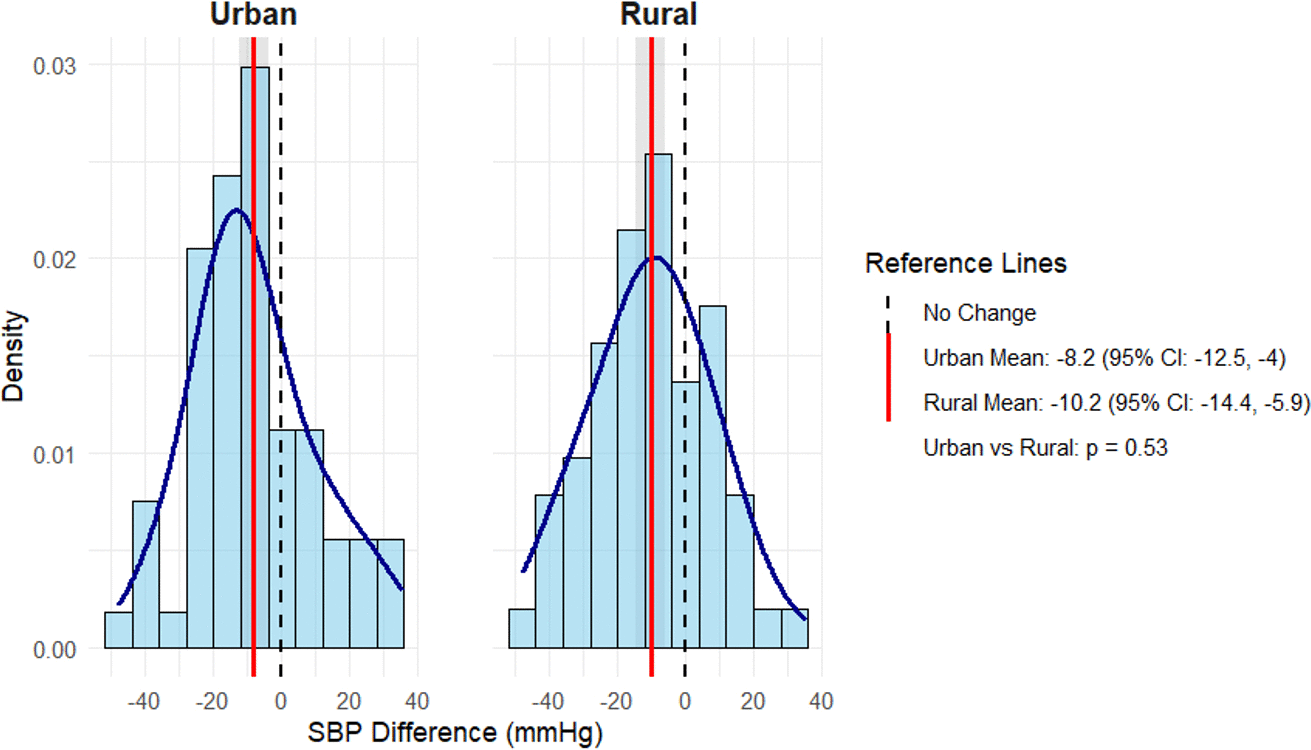
