## Supplementary Table 1 for "Community Blood Pressure groups – examining the acceptability and other effects of a pilot intervention in Zimbabwe"

**Supplementary Table 1:** Change in BMI from baseline to follow-up, overall and by residential area and sex.

| **Group/Category** | | **n** | **BMI decreased, n (%)** | ***p-value** |
| --- | --- | --- | --- | --- |
| **Overall** | | 137 | 76 (55.5) | 0.078 |
| **Residential Site** | **Urban** | 70 | 46 (65.7) | **0.012** |
|  | **Rural** | 67 | 30 (44.8) | 0.798 |
| **Sex** | **Male** | 29 | 13 (44.8) | 0.393 |
|  | **Female** | 108 | 63 (58.3) | **0.017** |

BMI, body mass index. P-values compare paired BMI measurements at baseline and follow-up using the Wilcoxon signed-rank test.
